# Personal protective equipment for reducing the risk of COVID-19 infection among healthcare workers involved in emergency trauma surgery during the pandemic: an umbrella review

**DOI:** 10.1101/2020.09.24.20201293

**Authors:** Dylan P Griswold, Andres Gempeler, Angelos Kolias, Peter J. Hutchinson, Andres Rubiano

**Author notes:** Correspondence: Andres Rubiano, Neuroscience Institute, INUB-MEDITECH Research Group, El Bosque University, Bogotá, Colombia.

## Abstract

**Objective:** The objective of this review was to summarise the effects of different personal protective equipment (PPE) for reducing the risk of COVID-19 infection in health personnel caring for patients undergoing trauma surgery. The purpose of the review was to inform recommendations for rational use of PPE for emergency surgery staff, particularly in low resources environments where PPE shortages and high costs are expected to hamper the safety of healthcare workers (HCWs) and affect the care of trauma patients.

**Introduction:** Many healthcare facilities in low-and middle-income countries are inadequately resourced. COVID-19 has the potential to decimate these already strained surgical healthcare services unless health systems take stringent measures to protect healthcare workers from viral exposure.

**Inclusion criteria:** This review included systematic reviews, experimental and observational studies evaluating the effect of different PPE on the risk of COVID-19 infection in HCWs involved in emergency trauma surgery. Indirect evidence from other healthcare settings was considered, as well as evidence from other viral outbreaks summarised and discussed for the COVID-19 pandemic.

**Methods:** We conducted searches in the L·OVE (Living OVerview of Evidence) platform for COVID-19, a system that performs automated regular searches in PubMed, Embase, Cochrane Central Register of Controlled Trials (CENTRAL), and over thirty other sources. The risk of bias assessment of the included studies was planned with the AMSTAR II tool for systematic reviews, the RoBII tool for randomised controlled trials, and the ROBINS-I tool for non-randomised studies. Data were extracted using a standardised data extraction tool and summarised narratively. The Grading of Recommendations, Assessment, Development, and Evaluation (GRADE) approach for grading the certainty of the evidence was followed.

**Results:** We identified 17 systematic reviews that fulfilled our selection criteria and were included for synthesis. We did not identify randomised controlled trials during COVID-19 or studies additional to those included in the reviews that discussed other similar viral respiratory illnesses.

**Conclusions:** The use of PPE drastically reduces the risk of COVID-19 compared with no mask use in HCWs in the hospital setting. N95 and N95 equivalent respirators provided more protection and were found to halve the risk of COVID-19 contagion in HCWs from moderate and high-risk environments. Eye protection also offers additional security and is associated with reduced incidence of contagion. These effects apply to emergency trauma care. Decontamination and reuse appear as feasible, cost-effective measures that would likely help overcome PPE shortages and enhance the allocation of limited resources.

**SUMMARY OF FINDINGS:** There is high certainty that the use of N95 respirators and surgical masks are associated with a reduced risk of coronaviruses respiratory illness when compared with no mask use. In moderate to high-risk environments, especially in aerosol-generating procedures, N95 respirators are associated with a more significant reduction in risk of COVID-19 infection compared with surgical masks. Eye protection also reduces the risk of contagion.

Decontamination of masks and respirators with ultraviolet germicidal irradiation, vaporous hydrogen peroxide, or dry heat is effective and does not affect PPE performance or fit.

***(Figure 1: GRADE summary of findings)***

## INTRODUCTION

Many healthcare facilities in low-and middle-income countries are inadequately resourced. COVID-19 has the potential to decimate these already strained surgical healthcare services unless health systems take stringent measures to protect healthcare workers (HCWs) from viral exposure. A recent study showed that 15.6% of confirmed COVID-19 patients are symptomatic and that nearly half of patients with no symptoms at detection time will develop symptoms later.^1^ Furthermore, the preoperative evaluation of emergency trauma patients is limited.

These factors impede and confound diagnostic triage. Improper infection prevention may create a ‘super-spreader’ event in a high-volume healthcare facility or reduce available personnel. Consequently, the infection control strategy of trauma surgery staff is a top priority.

To take care of patients, providers must first take care of themselves. Personal protective equipment (PPE) is paramount to protect health care workers from contracting the virus and becoming disease carriers. Basic recommended PPE for trauma surgery staff of high-income country facilities include: 1) a surgical mask or better for all personnel interacting with patients and in the OR (including cleaning staff); 2) N95 or better mask for all staff in close contact with the patients (<6 feet away); 3) PAPR for aerosolising and high-risk procedures (ear, nose, throat, thoracic, and transsphenoidal neurosurgery operations); 4) universal testing of patients pre-operatively to enable appropriate PPE use, and 5) changing scrubs after every procedure.^2^ These recommendations are suitable for high-resource settings but are less feasible in low-resource settings. A rapid-turnaround survey of 40 healthcare organisations across 15 LMICs revealed that 70% lack PPE and COVID-19 testing kits, and only 65% of the respondents showed confidence in hospital staff’s knowledge about precautions to be taken to prevent COVID-19 infection among hospital personnel.^3^

Some resource-adjusted recommendations include the use of cloth masks and bandanas. While innovative, their moisture retention, reusability, and filtration are considered very inferior to N95, and other masks.^4^ What is most needed is evidence guided recommendations for PPE use and COVID-19 screening in LMICs surgical systems where resources are either limited or unavailable. HCWs have been instructed to consider refraining from caring for patients in the absence of adequate PPE availability.

A preliminary search of PROSPERO, MEDLINE, the Cochrane Database of Systematic Reviews, and the *JBI Database of Systematic Reviews and Implementation Reports* was conducted, and no current or underway systematic reviews on the topic were identified.

The primary objective of the review was to summarise the effects of different personal protective equipment in reducing the risk of COVID-19 infection of health personnel caring for patients undergoing trauma surgery. The purpose of the review was to inform recommendations for the rational use of PPE in emergency surgery staff, particularly in low resources environments where PPE shortages and high costs are expected to hamper the safety of HCWs and affect the care of trauma patients. A preliminary search of PROSPERO, MEDLINE, the Cochrane Database of Systematic Reviews, and the JBI Database of Systematic Reviews and Implementation Reports was conducted, and no current or underway systematic reviews on the topic were identified.

## REVIEW QUESTIONS

We set to synthesise the available evidence on the effects of various personal protective equipment (PPE) in reducing the risk of COVID-19 infection of health personnel caring for patients undergoing trauma surgery. We were also interested in data on the costs associated with the use of PPE since it is a vital aspect to consider when generating recommendations for low-resources environments.

## INCLUSION CRITERIA

### Participants

We considered studies that included HCWs in emergency trauma surgery settings during the COVID-19 pandemic. Given the likelihood that reports on this specific population were scarce or even non-existent, we also included studies of HCWs in any procedural and in-hospital setting, such as the operating room, the emergency room, and critical care units. Furthermore, indirect evidence from other viral respiratory diseases (especially SARS and MERS) was considered if summarised and discussed regarding the COVID-19 pandemic.

### Intervention(s)

Different types of PPE used while caring for patients in hospital settings (preferably in emergency surgery).

### Comparator(s)

Comparators of interest were no PPE use and different types of PPE.

### Outcomes

The primary outcome of interest was the risk of contagion to health personnel involved in the care of the described population during the COVID-19 pandemic, expressed as incidence, or with association measures such as risk ratios or odds ratio when compared to different PPEs or no-PPE. We were also interested in summarising evidence of costs associated with PPE use during the pandemic.

### Types of studies

This review considered systematic reviews of experimental and observational studies, and experimental or observational studies if not included in systematic reviews that fulfilled population and intervention criteria. We also included reports of costs associated with the use of PPE and reports on implementation strategies that could inform recommendations for low resource settings. Only studies published in English or Spanish were included. We included preprint studies identified in our search, but no ongoing studies were considered.

## METHODS

We conducted a broad evidence synthesis (umbrella review) to summarise the effects of PPE on the risk of COVID-19 infection in healthcare workers (HCW) caring for patients in need of emergency surgery due to trauma. A protocol of this review following the PRISMA statement was registered in the International Prospective Registry of Systematic Reviews (PROSPERO; CRD42020198267). This review was conducted following the JBI methodology for systematic reviews of aetiology and risk.^5^

### Search strategy

We conducted searches in the L·OVE (Living OVerview of Evidence) platform for COVID-19. The platform was consulted on Jul 27 2020, using the entries: 1) Prevention or treatment - Procedures – Protective measures - PPE + Population Filter: COVID19; 2) Prevention or treatment - Procedures – Protective measures -PE + Population filter: Health workers.

### Information sources

The databases to be searched include the L·OVE (Living OVerview of Evidence) platform for COVID-19, a system that performs automated regular searches in PubMed, Embase, Cochrane Central Register of Controlled Trials (CENTRAL), and over thirty other sources. When compared to manual searches, this platform consistently identifies all the available studies associated with the terms of interest. It allows for a fast (automated) search that is easy to update - a crucial element given the urgent need to answer the research question rapidly and thoroughly.

### Study selection

Following the search, all identified citations were collated and uploaded into EndNoteX9 (Clarivate Analytics, PA, USA). The citations were then imported into JBI SUMARI for the review process. Two independent reviewers examined titles and abstracts for eligibility. Full-text review verified fulfilment of selection criteria. All decisions taken during screening were documented and are outlined in this report with a list of excluded studies. Any disagreements that arose between the reviewers were solved by consensus. The results of the search are presented in a Preferred Reporting Items for Systematic Reviews and Meta-analyses (PRISMA) flow diagram (Figure 2).^6^

**Figure 1.**
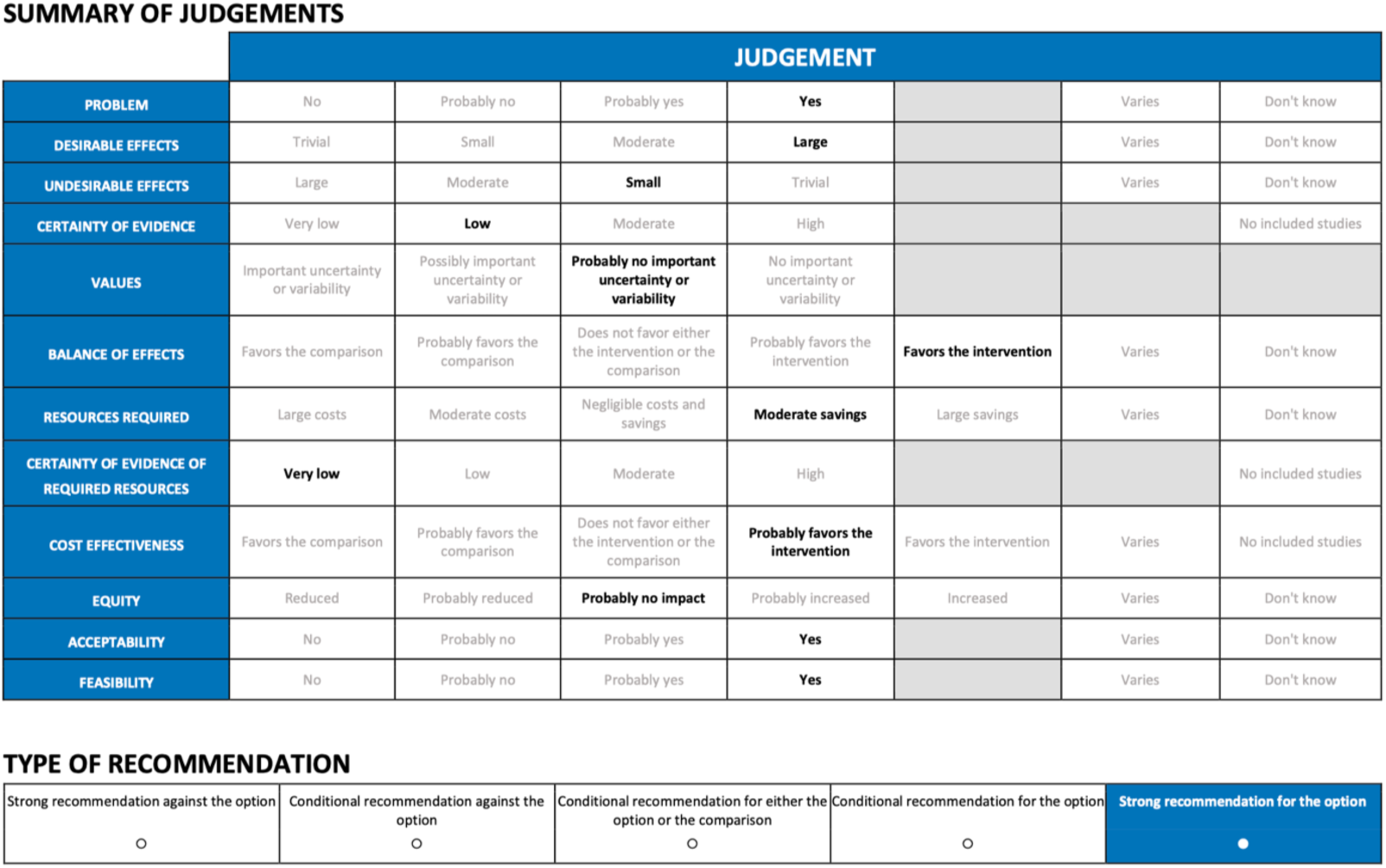
GRADE summary of judgements.

**Figure 2.**
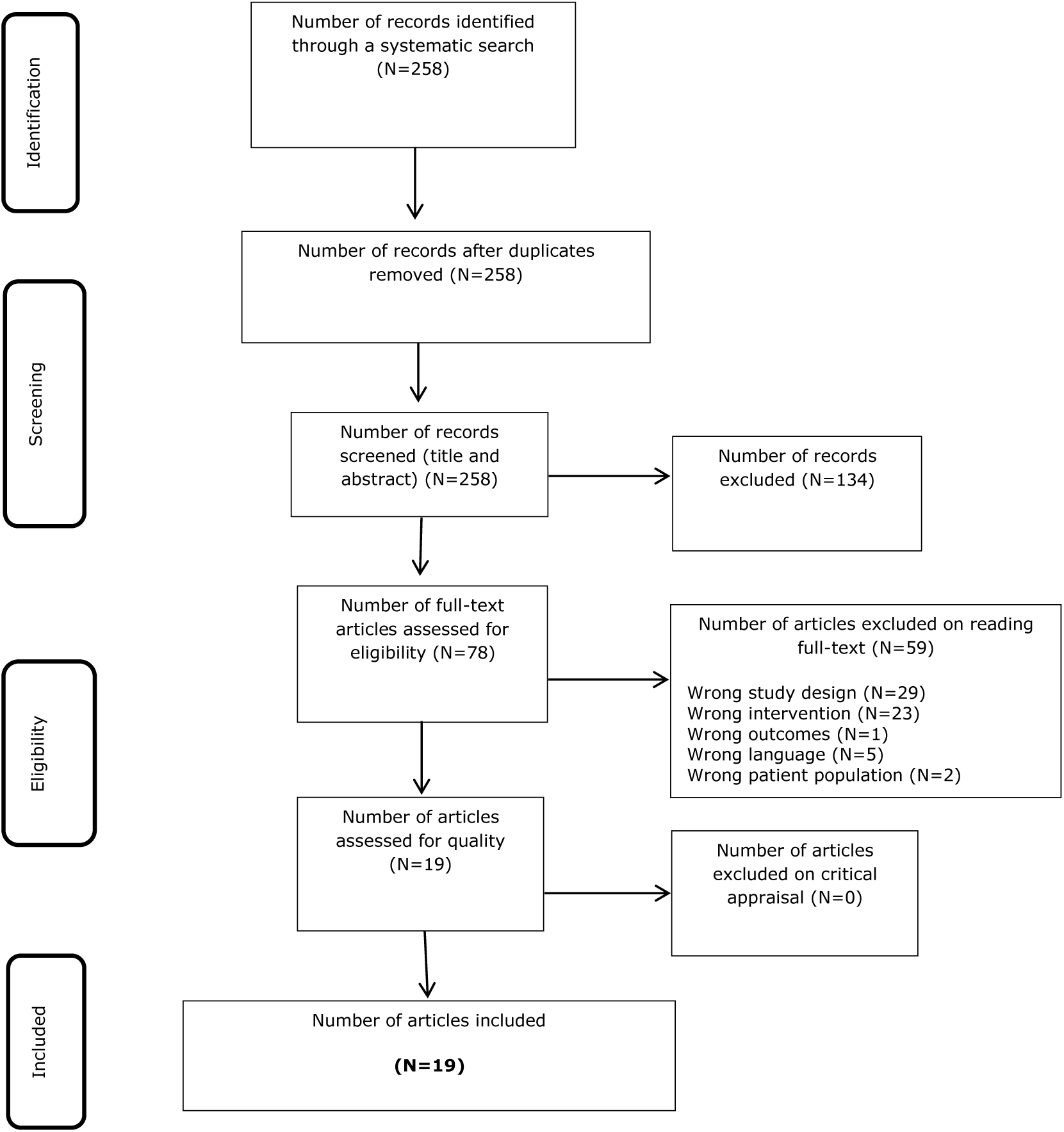
PRISMA search results, study selection and inclusion process.

### Assessment of methodological quality

Eligible studies were critically appraised by a reviewer and verified by a second reviewer using the AMSTAR tool. The risk of bias was assessed for only the primary outcome: infection of healthcare workers by COVID-19 or similar. The results of the critical appraisal are reported narratively and are considered for discussion of results.

All included studies, regardless of their risk of bias, underwent data extraction and synthesis.

### Data extraction

Data were extracted from the included studies by a reviewer and verified by a second reviewer using a data extraction tool from JBI SUMARI.^5^

The data extracted include specific details about the populations, study methods, interventions, and outcomes of significance to the review question and specific objectives. Disagreements were solved by consensus.

### Data synthesis

Studies were summarized narratively considering their scope, number of included studies, and risk of bias. Effect sizes from systematic reviews and individual studies not included in them are expressed as odds ratios (for dichotomous data) with their 95% confidence intervals.

### Assessing certainty in the findings

The Grading of Recommendations, Assessment, Development, and Evaluation (GRADE) approach for grading the certainty of the evidence was followed. Grading the certainty of the evidence was not undertaken if adaptation from the identified reviews using the GRADE approach was considered complete and adequate.^7,8^ The certainty of the evidence was considered for interpretation and discussion of findings.

## RESULTS

### Study inclusion

The study selection process is illustrated in Figure 1.^6^ The described search identified a total of 258 records. After title and abstract screening, 78 studies were considered for full-text review, of which 59 were excluded. Reasons for exclusion were: wrong study design (n=29), ^9–37^ wrong intervention (n=23),^38–59^ wrong outcomes (n=1),^60^ wrong language (n=5),^61–65^ wrong patient population (n=2).^66,67^ This left 19 studies for appraisal, extraction, and synthesis.^4,68–83^

Appendix I shows a list of the 59 excluded studies, with reasons for their exclusion. In general, studies were excluded because they had a wrong study design (mostly narrative reviews, guideline recommendations, and case reports), wrong setting (non-hospital settings, community setting), and not reporting data related to COVID-19 contagion in healthcare workers.

### Methodological quality

Tables 1-3 show the results of the critical appraisal of the methodological quality. Overall, the quality of the 19 included studies was assessed as moderate to high by JBI appraisal standards, and no disagreements occurred between the reviewers. Of the 17 included systematic reviews, nine fulfilled all 11 indicators of the critical appraisal tool,^69,70,72,75–77,79,81,82^ one fulfilled ten indicators,^68^ choosing not to perform risk of bias assessment given the rapid publication of the review; four fulfilled nine indicators,^4,80,83,84^ failing to report a risk of bias assessment and choosing not to combine studies for meta-analysis owing to study limitations and heterogeneity in study designs, comparisons, and analyses. Two fulfilled six indicators,^71,78^ having no method of study appraisal, no method of minimising errors in data extraction, failing to report a risk of bias assessment, and choosing not to combine studies for meta-analysis owing to study limitations and heterogeneity in study designs, comparisons, and analyses; and one fulfilled four indicators,^73^ for not reporting the indicators aforementioned in the previous studies in addition to a lack of future directives and recommendations for policy and clinical practice. All systematic reviews clearly stated the review question, applied appropriate inclusion criteria and search strategy.

### Critical Appraisal Results

**Table 1:** Case Series.

| Citation | Q1 | Q2 | Q3 | Q4 | Q5 | Q6 | Q7 | Q8 | Q9 | Q10 |
| --- | --- | --- | --- | --- | --- | --- | --- | --- | --- | --- |
| Wilson JM, Schwartz AM, Farley KX, Devito DP, Fletcher ND. 2020. | Y | Y | Y | Y | Y | Y | Y | Y | Y | Y |
| % | 100.0 | 100.0 | 100.0 | 100.0 | 100.0 | 100.0 | 100.0 | 100.0 | 100.0 | 100.0 |

**Table 2:** Qualitative Research.

| Citation | Q1 | Q2 | Q3 | Q4 | Q5 | Q6 | Q7 | Q8 | Q9 | Q10 |
| --- | --- | --- | --- | --- | --- | --- | --- | --- | --- | --- |
| Houghton C, Meskell P, Delaney H, Smalle M, Glenton C, Booth A, et al. 2020. | Y | Y | Y | Y | Y | Y | Y | Y | Y | Y |
| % | 100.0 | 100.0 | 100.0 | 100.0 | 100.0 | 100.0 | 100.0 | 100.0 | 100.0 | 100.0 |

**Table 3:**
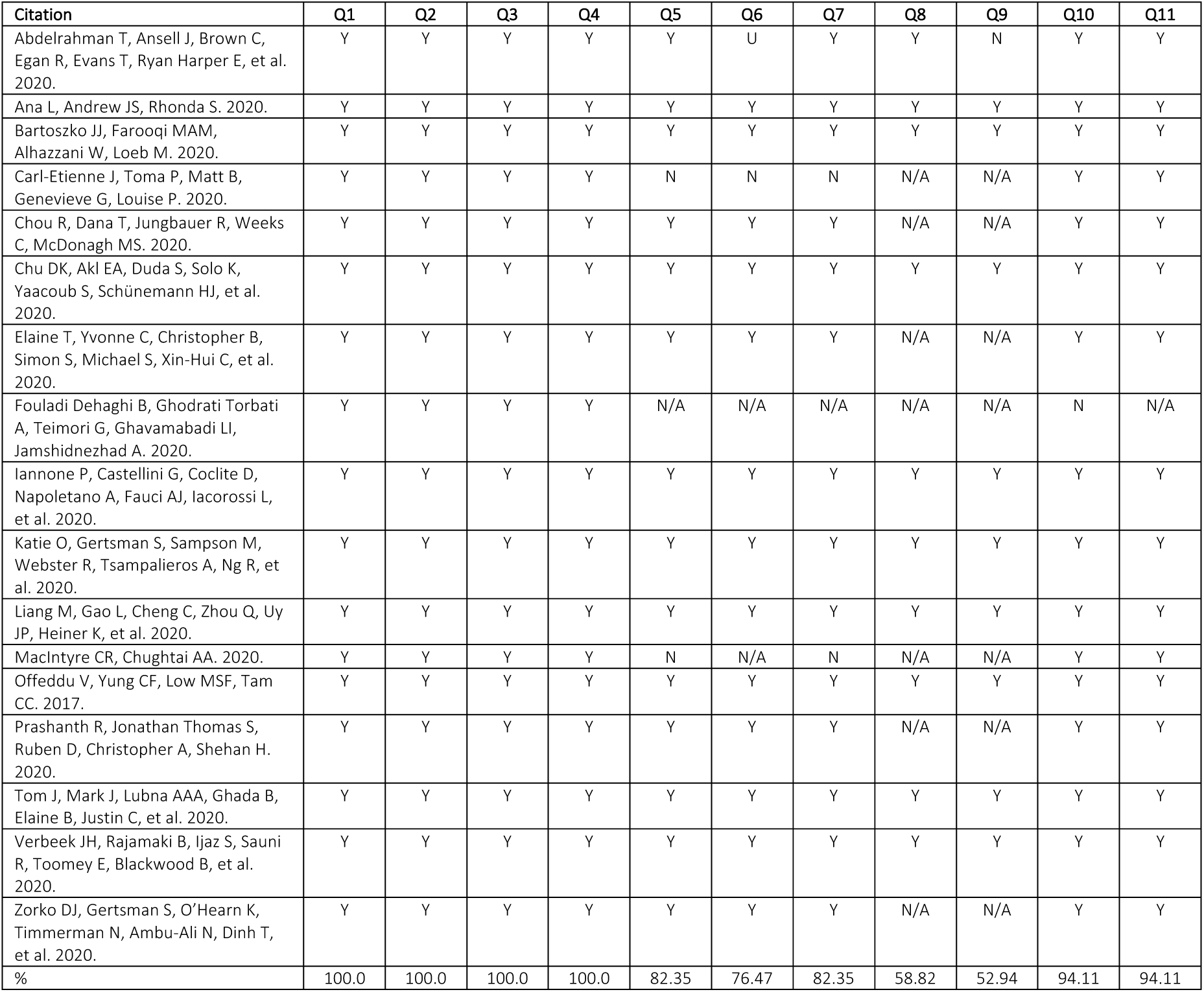
Systematic Reviews.

### Characteristics of included studies

The 17 included studies were systematic reviews, ^4,68–73,75,76,79–84^. Appendix II provides details of the characteristics of the included studies. All but one study was published in 2020.^79^ Data extracted from reviews included thousands of participants from 35 different countries.

Ten of 17 systematic reviews evaluated the risk of contagion for respiratory viral infections,^4,35,69–72,77,82^ which six included outcome data for COVID-19 infection.^4,69,72,75,77,78,82^

Four systematic reviews,^70,75,79,81^ evaluate other respiratory pathogens such as seasonal influenza, SARS, H1N1, and MERS.

### Review findings

We did not identify comparative studies of PPE effect on the risk of COVID-19 contagion in the emergency surgery setting. We did identify observational studies of COVID-19 in HCW, as well as experimental and observational studies that also addressed this question in HCWs regarding other coronavirus epidemics (SARS and MERS epidemics) considered generalisable to the COVID-19 pandemic. Some studies also assessed and summarised evidence from other viral respiratory illnesses, such as H1N1 or influenza, and reported results consistent with those of the coronaviruses outbreaks.

A high-quality systematic review that evaluated the effect of physical distancing face masks and eye protection on preventing COVID-19 contagion included 172 studies, considering evidence from COVID-19, MERS, and SARS.^72^ The authors identified 30 comparative studies that focused on the effect of different masks and respirators on virus transmission in healthcare workers or patients and 13 studies that addressed the same effect for eye protection.

They report that the use of a surgical mask compared with no face mask was associated with a considerable reduction in risk of contagion (OR= 0.33, 95% CI= 0.17–0.61). An even larger effect was seen when comparing N95 and N95 equivalent respirators to no mask (OR= 0.04, 95% CI= 0.004–0.30). Such estimates are based on studies, including a total of 12,817 participants. Adjusted and unadjusted studies were considered, and both estimates were consistent with the mentioned effect on contagion risk reduction when considering N95 or surgical/medical masks vs. no mask (adjusted OR= 0.15 (0.07 to 0.34); unadjusted RR 0.34 (95% CI 0.26 to 0.45). Evidence for the precisely estimated reduction was rated as low by the authors, given some inconsistency and risk of bias. Nevertheless, the beneficial effect of mask protection was large, and they considered it of high certainty.^72^ They report that N95 had a stronger protective association compared with surgical masks or 12–16-layer cotton masks, and both N95 and surgical masks also had a stronger association with protection versus single-layer masks.

Regarding the use of eye protection, pooled analysis of 13 unadjusted and two adjusted studies suggested a reduced risk of contagion with eye protection compared with no eye protection (unadjusted: RR= 0.34, 95% CI 0.22 to 0.52; adjusted OR= 0.22, 95% CI 0.12 to 0.39). This review was considered pivotal due to the high number of included studies, the recent date of publication, and the adequacy of methods and reporting. Challenges reported in the studies included frequent discomfort, high resource use, less clear communication, and perceived reduced empathy of care providers by their patients.^72^

A rapid systematic review that also addressed the effect of masks to prevent COVID-19 infection considered evidence from the current pandemic in addition to the SARS and MERS epidemics.^4^The review reports a reduction of risk of transmission associated with the use of masks in general. It suggests a more significant reduction associated with N95 respirators compared to surgical masks in the hospital setting (an effect seen for COVID-19 independently, as well as with the other coronaviruses outbreaks).

Other reviews considered evidence from viral respiratory illnesses, including influenza or H1N1, and report a beneficial effect of PPE (medical masks or N95 respirators) on contagion risk reduction. ^70,75,77–79,81^ One of these reviews reports that the use of masks by HCWs and non-HCWs can reduce the risk of respiratory virus infection by 80% compared to no-mask (OR = 0.20, 95% CI = 0.11–0.37).^77^ Furthermore, respirators were found to be more protective than surgical masks; and surgical masks more protective than cloth masks.^78^ There appears to be no difference between respirators and medical masks when used in non-aerosol generating procedures low-risk environments)^70,81^ Conversely, no significant evidence was found that supported an equivalence claim of medical masks with respirators in their level of protection against COVID-19 or other similar viruses.^80^ In moderate and high-risk hospital settings, N95 are associated with more significant reductions in risk of contagion.^4,78^

A systematic review based on experimental designs only found that N95 respirators halve the risk of any respiratory illness compared to surgical masks; the certainty of the evidence was low due to baseline differences, indirectness of evidence for COVID-19, and low event rates that account for imprecision.^75^ The reduction in contagion risk calculated from 2 RCTs was estimated to be: RR 0.43, 95% CI 0.29, 0.64; I2 = 0%, from pooled analysis; with an absolute effect of preventing 73 (95% CI= 91 - 46) more infections per 1000 HCWs wearing N95 respirators compared with surgical masks.^75^

Among the included studies, one reported on the use of Powered Air Purifier Respirators (PAPR).^69^Based on observational studies, the authors report they did not found a difference in risk of contagion in HCWs when comparing PAPR devices with other, more compliant protective elements (N95, FFP2). They found that PAPR users reported higher heat tolerance but limited mobility and reduced audibility.

Regarding decontamination, we included a systematic review that assessed the effectiveness of ultra-violet germicidal irradiation (UVGI) for the decontamination of PPE and its impact on PPE performance.^76^ Their findings support that the use of a cumulative UV-C dose of at least 40,000 J/m2 results in adequate decontamination without affecting performance or fit afterwards. Another review on the subject reported that mask (N95) performance was best conserved using dry heat decontamination, and that vaporous hydrogen peroxide, as well as UVGI, are effective decontaminants. However, its effect on surgical masks is unknown.^83^ The authors also state that bleach is not safe for decontamination since it alters mask performance and might be associated with health risk for users.

A systematic review that searched for barriers and facilitators of HCWs adherence to PPE protocols included 20 studies of moderate to high-quality overall (10 from Asia, four from Africa, four from Central and North America, and two from Australia).^74^ They report that HCWs were unsure to follow recommendations when they are long and ambiguous or do not reflect national or international guidelines. Some were overwhelmed because of constantly changing guidelines and by the increased workload and fatigue associated with PPE use due to preparation and cleaning. A serious concern was the lack of PPE or the low quality of the available items, pointing at a need to adjust supplies during the pandemic. HCWs reported that it was challenging to use masks and other equipment when it made patients feel isolated, frightened, or stigmatised. Of course, discomfort associated with wearing PPE was also reported.

## Discussion

Our review aimed at summarising the available evidence of the effect of different PPE on the risk of COVID-19 infection among HCWs caring for patients requiring urgent trauma assessment and surgical care. We did not find experimental studies that assessed PPE on emergency trauma surgery settings during the pandemic. Limited observational evidence from COVID-19, indirect evidence from other healthcare settings, and other viral outbreaks were all considered to answer our research question given that the population (HCW) and intervention (PPE) of interest were the same and thus considered applicable to the emergency surgery setting.

The available evidence was consistent to show that the use of N95 respirators and surgical masks is associated with a reduced risk of coronaviruses respiratory illness compared with no mask use, with high certainty on this beneficial effect.^4,72^ In moderate to high-risk environments, especially in aerosol-generating procedures, evidence suggests that N95 respirators are associated with a more significant reduction in risk of COVID-19 infection compared with surgical masks; an effect seen in observational COVID-19 studies and experimental viral respiratory illness studies. Low-quality evidence estimates from these studies suggest a relative reduction of 50% in the risk of contagion associated with N95 respirators compared to surgical masks. Eye protection also significantly reduces the risk of contagion compared to no-eye protection. Furthermore, the decontamination of masks and respirators with ultraviolet germicidal irradiation, vaporous hydrogen peroxide, or dry heat is effective and does not affect PPE performance or fit. This evidence should inform decontamination and reuse protocols to avoid shortages and enhance resource allocation and use.

The costs associated with additional protective measures during the COVID-19 pandemic could be significant and affect healthcare institutions in low and middle-income countries. The cost-effectiveness of interventions must also be taken into consideration to generate recommendations during the current pandemic. The possibility to decontaminate and reuse different types of masks can be determinant in shortages and will probably reduce costs without affecting HCW’s safety. A cost analysis study compared the use of disposable FFP3 standard masks vs. SR

100 reusable respirators.^85^ Disposable masks are indicated to be replaced after each surgical case, and each one costs roughly $4.27 (USD). The cost per unit for a reusable respirator, supplied with an appropriate filter, is approximately $44(USD), and replacement filters cost $0.37. The authors of this economic evaluation highlighted that reusable PPE could be associated with considerable cost savings and estimated that the cost of acquiring a respirator is recovered after it is used for the care of 10 patients.^85^

In a survey of 5,442 neurosurgical staff members in Hubei province, among 120 participants that were infected, 78.3% reported wearing surgical masks, and 20.8% failed to use any protection when exposed to the source of infection. A total of 1,287 operated under level 2 protection, and only one was infected,^29^ further illustrating the pertinence of wearing adequate PPE when caring for surgical trauma patients.

Expert recommendations developed from a study of emergency tracheal intubation in 202 patients with COVID-19 in Wuhan, China, notes that while PAPRs were the PPE of choice when face shields or full hoods without PAPR were substituted, there were no instances of infection of operators.^10^ As the risk of virus exposure due to self-contamination is high during the removal of PPE, educational training for proper donning and doffing of PPE as well as monitoring for compliance is essential. The minimum recommended PPE is eye protection, a fit-tested respirator (N95 or FFP3), a fluid-resistant gown, and gloves. The French guidelines recommend FFP2-type protective filtering masks when performing any aerosol-generating procedures.^12^ Guidelines for chest compressions recommend ‘level three’ PPE, which includes an FFP, disposable fluid-resistant gown, disposable apron, and gloves, fluid-resistant surgical mask, and eye or full-face protection.^17^ Recommendations for otolaryngologists include fluid-resistant FFP3/N95 mask, disposable and fluid resistant gloves, and gown, glasses, or a full face shield.^14^

Recommendations for safe orthopaedic surgery practices state that surgery should only be performed on COVID-19 positive patients when the risks of surgery are outweighed by the benefits, such as in emergencies or when a surgical delay could cause increased morbidity and mortality.^11^ The recommended PPE are N95 respirators or PAPR with the use of face shields, isolation gowns, boot covers, and gloves.

Intubation of trauma patients is a high-risk-of-contagion procedure during the COVID-19 pandemic. A survey in 503 hospitals from 17 countries included 1,718 HCWs performing 5,148 tracheal intubations and measured a 10.7% incidence of COVID-19 infection after tracheal intubation.^32^ Most participants reported wearing gloves, gown, eye protection, and FFP2/FFP3/N95/N100 respirators. Simulation studies have assessed the effect of additional protective and preventive measures, such as transparent plastic boxes or PAPR, on the vision, comfort, and success of tracheal intubations.^35,36^

A survey of HCWs realities and perceptions during the pandemic in Latin America included 936 participants and reported low access to disposable gowns (67.3%), N95 respirators (56.1%), and facial protective shields (32.6%). Even access to disposable surgical masks was reported by only 83.9% of participants.^33^ This emphasises the need for rational use of limited PPE during the pandemic in LMICs to ensure HCW safety without withholding urgent trauma care.

Our findings regarding decontamination should be considered as a feasible solution for the limited access to N95 equivalent respirators during shortages and in limited resources environments. Also, to avoid such shortages, it appears that N95 respirator equivalents use should be limited to moderate to high-risk environments; when caring for patients with confirmed COVID-19, or for suspicious or unknown status patients that need emergency surgery due to trauma.

### Strengths and Limitations

Our review employed an automated search platform where evidence on COVID-19 is available. This strategy streamlined the rapid nature of the review while ensuring that all relevant studies were identified. Using an automated system has the additional long-term advantage of facilitating review updates by quickly identifying new studies that satisfy selection criteria. Our review also has the strength of having critically assessed all included studies. We report on the estimates and evidence grading of the identified high-quality systematic reviews. A metanalysis of systematic reviews results was not planned in our review protocol and was not considered adequate, given the overlap of included studies among reviews and the variation in selection criteria. Despite the differences between reviews, the consistency of findings among the reviews provides high certainty of the beneficial effects of PPE in the hospital setting. The main limitation of our review is that evidence from our specific setting of interest - emergency trauma surgery - was not identified. Nevertheless, extrapolation from other clinical settings such as the emergency room, COVID-19 wards and critical care during the pandemic was considered adequate given the characteristics of the intervention and the similarities to the setting of interest.

## Conclusions

The use of PPE drastically reduces the risk of COVID-19 compared with no mask use in HCWs in the hospital setting. Respirators like N95 or equivalent provided more protection and were found to halve the risk of COVID-19 contagion in HCWs from moderate and high-risk settings. Eye protection also provides additional protection and is associated with reduced incidence of contagion. These effects apply to emergency trauma care. Decontamination and reuse appear as feasible, cost-effective measures than could help overcome PPE shortages and enhance the allocation of limited resources.

### Recommendations for practice

When caring for a trauma patient with suspected or unknown COVID-19 status, HCWs should use at least N95 respirators or equivalents to reduce the risk of COVID-19 infection adequately. Decontamination with ultraviolet light, hydrogen peroxide, and dry heat should be made available.

### Recommendations for research

Robust RCTs comparing the efficacy of surgical masks vs N95 respirators in HCWs caring for trauma patients are potentially unethical, as existing data show a significant protective effect, thus requiring emergency trauma surgery staff to wear N95 respirators when available. As of October 2020, there is a lack of consensus among international experts surrounding the topic of aerosol transmission, meaning that viral micro-droplets are capable of floating in the air without being pulled down by gravity. This means that if someone coughs, sings, or even breathes, the micro-droplets can stay in stagnant air for up to 16 hours, and with normal ventilation between 20 minutes to four hours. While multiple studies have discussed how SARS-CoV-2 can be found in aerosols, including one from May and another from April,^86,87^ a group of epidemiologists in late July characterised research on aerosol transmission as unconvincing and cited extensive published evidence from across the globe showing the overwhelming majority of viral spread is via large respiratory droplets.^88^ The CDC did not acknowledge aerosol transmission as an important route for viral transmission until September 2020, placing aerosol ahead of droplet transmission as the predominant mechanism of viral spread. However, just a few days later the statement was recalled, with updated guidelines saying HCWs need an N95 respirator for aerosol-generating procedures, only.^89^ The hospital administrators and epidemiologists who argue that the virus is mainly droplet-spread claim N95 respirators and strict patient isolation practices are not necessary for routine care of COVID-19 patients. It is essential to develop a complete understanding of the transmissibility of SARS-CoV-2 as it drives two different sets of protective practices, touching on everything from airflow within hospital wards to patient isolation to choices of PPE. Enhanced protections would be expensive and disruptive and would have strong implications on cost-effectiveness data, especially for low-income environments. Amid the uncertainty, adopting the highest possible forms of protection seems the best course of action.

(Figure 3: GRADE conclusions)

**Figure 3.**
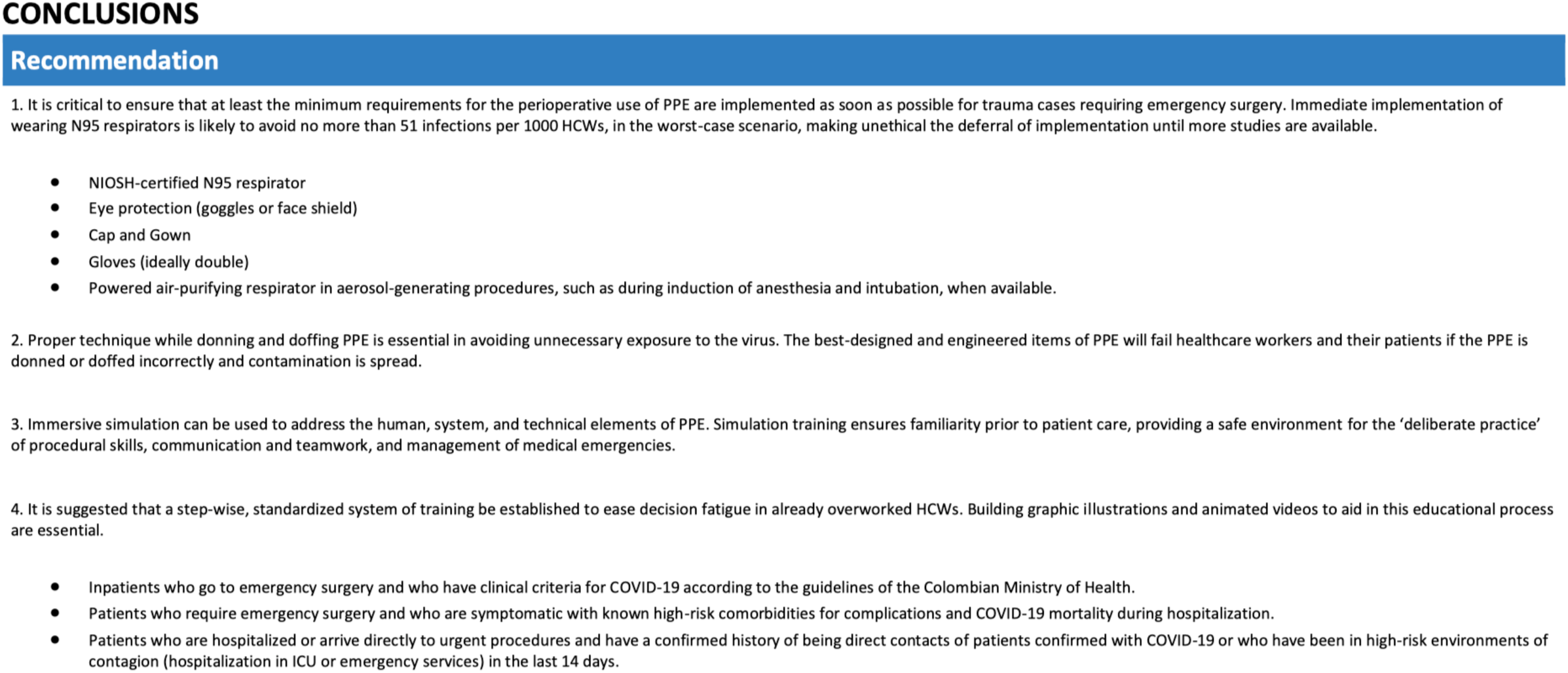
GRADE recommendations and conclusions.

## Data Availability

All data is available upon request

## Acknowledgements

None to report

## APPENDICES

Appendix I: Studies excluded on full text

**Parreira PCL et al. Personal protective masks for COVID-19 prevention: quick systematic review**.

Reason for exclusion: Portuguese language

**Schnitzbauer AA et al. SARS-CoV-2/COVID-19: systematic review of requirements for personal protective equipment in primary patient contact and organisation of the operating area**.

Reason for exclusion: German language

**Comité Provincial de Biotecnología. Use of face masks in the context of the COVID-19 pandemic in the Neuquén Health System**.

Reason for exclusion: Spanish language

**ANVISA. Guidelines for health services: prevention and control measures that should be adopted when assisting suspected or confirmed cases of infection with the new coronavirus (SARS-CoV-2)**.

Reason for exclusion: Portuguese language

**Instituto de Evaluación de Tecnologías en Salud e Investigación. Comparison of surgical (medical) masks with respirators to prevent SARS-COV-2 infection in health personnel to COVID-19**.

Reason for exclusion: Spanish language

**Gralton et al. Protecting healthcare workers from pandemic influenza: N95 or surgical masks?**

Reason for exclusion: Wrong patient population: healthcare workers at high risk for occupationally acquired influenza.

**Brainard J et al. Facemasks and similar barriers to prevent respiratory illness such as COVID-19: A rapid systematic review**.

Reason for exclusion: Wrong patient population: influenza-like illness in community settings

**Canova V et al. Transmission risk of SARS-CoV-2 to healthcare workers - observational results of a primary care hospital contact tracing**.

Reason for exclusion: wrong outcome: no comparable outcome data for risk of contagion

**Dedeilia et al. Medical and Surgical Education Challenges and Innovations in the COVID-19 Era: A Systematic Review**. Reason for exclusion: wrong intervention: the review aimed to identify the challenges imposed on medical and surgical education by the COVID-19 pandemic and the proposed innovations enabling the continuation of medical student and resident training.

**Jones P et al. What proportion of healthcare worker masks carry virus? A systematic review**.

Reason for exclusion: wrong intervention: this review aimed to determine the carriage of respiratory viruses on facemasks used by HCW

**Begg S et al. Can we use these masks? Rapid Assessment of the Inhalation Resistance Performance of Uncertified Medical Face Masks in the Context of Restricted Resources Imposed during a Public Health Emergency**.

Reason for exclusion: Wrong intervention: assessment of filtration performance of surgical masks

**Malysz M et al. Resuscitation of the patient with suspected/confirmed COVID-19 when wearing personal protective equipment: A randomised multicenter crossover simulation trial**.

Reason for exclusion: Wrong intervention: median chest compression depth and rate

**Islam MS et al. Examining the current intelligence on COVID-19 and infection prevention and control strategies in health settings: A global analysis**.

Reason for exclusion: Wrong intervention: transmission dynamics and pathogenic and clinical features of COVID-19

**Cadnum JL et al. Effectiveness of Ultraviolet-C Light and a High-Level Disinfection Cabinet for Decontamination of N95 Respirators**.

Reason for exclusion: Wrong intervention: evaluating decontamination methods for N95 respirators

**Zulauf KE et al. Microwave-Generated Steam Decontamination of N95 Respirators Utilising Universally Accessible Materials**.

Reason for exclusion: Wrong intervention: evaluating decontamination methods for N95 respirators

**Zhong H et al. Reusable and Recyclable Graphene Masks with Outstanding Superhydrophobic and Photothermal Performances**.

Reason for exclusion: Wrong intervention: evaluating decontamination methods for N95 respirators

**Jones P et al. What proportion of healthcare worker masks carry virus? A systematic review**.

Reason for exclusion: Wrong intervention: This review aimed to determine the carriage of respiratory viruses on facemasks used by HCW.

**Derr TH et al. Aerosolized Hydrogen Peroxide Decontamination of N95 Respirators, with Fit-Testing and Virologic Confirmation of Suitability for Re-Use During the COVID-19 Pandemic**.

Reason for exclusion: Wrong intervention: evaluating decontamination methods for N95 respirators

**Christensen L et al. A randomised trial of instructor-led training versus video lesson in training health care providers in proper donning and doffing of personal protective equipment**.

Reason for exclusion: wrong intervention: evaluated method of education for donning and doffing

**Leormandi R et al. Effect of ethanol cleaning on the permeability of FFP2 mask**.

Reason for exclusion: Wrong intervention: evaluating decontamination methods for N95 respirators

**Avilash C et al. Analysis of SteraMist ionised hydrogen peroxide technology as a method for sterilising N95 respirators and other personal protective equipment**.

Reason for exclusion: Wrong intervention: evaluating decontamination methods for N95 respirators

**Massey T et al. Quantitative form and fit of N95 filtering facepiece respirators are retained after dry and humid heat treatments for coronavirus deactivation**.

Reason for exclusion: Wrong intervention: evaluating form and fit after decontamination methods for N95 respirators

**Cramer A et al. Disposable N95 Masks Pass Qualitative Fit-Test But Have Decreased Filtration Efficiency after Cobalt-60 Gamma Irradiation**.

Reason for exclusion: Wrong intervention: evaluating form and fit after decontamination methods for N95 respirators

**Simmons S et al. Disinfection effect of pulsed xenon ultraviolet irradiation on SARS-CoV-2 and implications for environmental risk of COVID-19 transmission**.

Reason for exclusion: Wrong intervention: evaluating decontamination methods to prevent COVID-19 transmission

**Simeon C et al. Thermal Disinfection Inactivates SARS-CoV-2 in N95 Respirators while Maintaining Their Protective Function**.

Reason for exclusion: Wrong intervention: evaluating decontamination methods for N95 respirators

**Ebru O et al. Vapor H2O2 sterilisation as a decontamination method for the reuse of N95 respirators in the COVID-19 emergency**.

Reason for exclusion: Wrong intervention: evaluating decontamination methods for N95 respirators

**Derince T. Effect of Using Barrier Devices for Intubation in COVID-19 Patients**.

Reason for exclusion: Wrong intervention: testing barrier devices for intubation

**Mhango M et al. COVID-19 Risk Factors Among Health Workers: A Rapid Review**.

Reason for exclusion: Wrong intervention: This study evaluated evidence on Covid-19 risk factors among HCWs

**Rubin GD et al. The Role of Chest Imaging in Patient Management during the COVID-19 Pandemic: A Multinational Consensus Statement from the Fleischner Society**.

Reason for exclusion: Wrong intervention: CT imaging

**Wahidi MM et al. The Use of Bronchoscopy during the COVID-19 Pandemic: CHEST/AABIP Guideline and Expert Panel Report**.

Reason for exclusion: Wrong study design: guidelines on the use of bronchoscopy during the COVID-19 pandemic

**Powell-Jackson T et al. Infection prevention and control compliance in Tanzanian outpatient facilities: a cross-sectional study with implications for the control of COVID-19**.

Reason for exclusion: Wrong intervention: Infection prevention and control compliance

**Liao L et al. Can N95 Respirators Be Reused after Disinfection? How Many Times?**

Reason for exclusion: Wrong intervention: investigated multiple commonly used disinfection schemes on media with a particle filtration efficiency of 95%.

**Konda A et al. Aerosol Filtration Efficiency of Common Fabrics Used in Respiratory Cloth Masks**.

Reason for exclusion: wrong intervention: filtration efficiency of common fabrics

**Qiangping et al. Epidemiological characteristics of COVID-19 in medical staff members of neurosurgery departments in Hubei province: A multicentre descriptive study**.

Reason for exclusion: Wrong study design: survey

**Ippolito M et al. Medical masks and Respirators for the Protection of Healthcare Workers from SARS-CoV-2 and other viruses**.

Reason for exclusion: wrong study design: narrative summary of surgical mask and respirator characteristics

**Yao W et al. Emergency tracheal intubation in 202 patients with COVID-19 in Wuhan, China: lessons learnt and international expert recommendations**.

Reason for exclusion: wrong study design: guideline recommendations for emergency tracheal intubation

**Service BC et al. Medically Necessary Orthopaedic Surgery During the COVID-19 Pandemic: Safe Surgical Practices and a Classification to Guide Treatment**.

Reason for exclusion: wrong study design: guideline recommendations for safe orthopaedic surgical practices during COVID-19 pandemic

**Lepelletier D et al. What face mask for what use in the context of COVID-19 pandemic? The French guidelines**.

Reason for exclusion: wrong study design: guideline recommendations for the use of face masks during COVID-19

**Leboulanger N et al. COVID-19 and ENT Pediatric otolaryngology during the COVID-19 pandemic. Guidelines of the French Association of Pediatric Otorhinolaryngology (AFOP) and French Society of Otorhinolaryngology (SFORL)**. Reason for exclusion: wrong study design: guideline recommendations for ENT pediatric otolaryngology during the COVID-19 pandemic

**Krajewska J et al. COVID-19 in otolaryngologist practice: a review of current knowledge**.

Reason for exclusion: wrong study design: Narrative review of the current knowledge on COVID-19-related information useful in otolaryngologist practice.

**Hirschmann MT et al. COVID-19 coronavirus: recommended personal protective equipment for the orthopaedic and trauma surgeon**.

Reason for exclusion: wrong study design: guideline recommendations for safe orthopaedic surgical practices during COVID-19 pandemic

**Godoy et al. Facial protection for healthcare workers during pandemics: a scoping review**.

Reason for exclusion: wrong study design: scoping review

**Brown E et al. Should chest compressions be considered an aerosol-generating procedure? A literature review in response to recent guidelines on personal protective equipment for patients with suspected COVID-19**.

Reason for exclusion: wrong study design: guideline recommendations on the role of chest compressions for patients with suspected COVID-19

**Malik T. COVID-19 and the Efficacy of Different Types of Respiratory Protective Equipment Used by Health Care Providers in a Health Care Setting**.

Reason for exclusion: wrong study design: case report

**Chersich MF et al. COVID-19 in Africa: care and protection for frontline healthcare workers**.

Reason for exclusion: wrong study design: recommendations

**Hiramatsu M et al. Anesthetic and surgical management of tracheostomy in a patient with COVID-19**.

Reason for exclusion: wrong study design: recommendations

**Lee S et al. Asymptomatic carriage and transmission of SARS-CoV-2: What do we know?**

Reason for exclusion: wrong study design: narrative recommendations

**Mueller AV et al. Assessment of Fabric Masks as Alternatives to Standard Surgical Masks in Terms of Particle Filtration Efficiency**.

Reason for exclusion: wrong study design: narrative review and recommendations regarding fabric masks in place of surgical masks

**Winck JC et al. COVID-19 pandemic and non invasive respiratory management: Every Goliath needs a David. An evidence based evaluation of problems**.

Reason for exclusion: wrong study design: narrative review describes some problems with the management of Covid-19 induced acute respiratory failure (ARF) by pulmonologists.

**Boskoski I et al. COVID-19 pandemic and personal protective equipment shortage: protective efficacy comparing masks and scientific methods for respirator reuse**.

Reason for exclusion: wrong study design: guideline recommendations on rational use and successful reuse of respirators

**Convissar D et al. Personal Protective Equipment N95 Facemask Shortage Quick Fix: The Modified Airway From VEntilatoR Circuit (MAVerIC)**.

Reason for exclusion: wrong study design: technical report proposes a makeshift filter mask

**Sugrue M et al. A cloth mask for under-resourced healthcare settings in the COVID19 pandemic**.

Reason for exclusion: wrong study design: narrative recommendations

**Jessop ZM et al. Personal Protective Equipment (PPE) for Surgeons during COVID-19 Pandemic: A Systematic Review of Availability, Usage, and Rationing**.

Reason for exclusion: wrong study design: guideline recommendations on the rational use of PPE for surgeons during COVID-19 pandemic

**Heinzerling A et al. Transmission of COVID-19 to Health Care Personnel During Exposures to a Hospitalised Patient - Solano County, California, February 2020**.

Reason for exclusion: wrong study design: anecdotal and descriptive

**Delgado D et al. Personal Safety during the COVID-19 Pandemic: Realities and Perspectives of Healthcare Workers in Latin America**.

Reason for exclusion: wrong study design: questionnaire

**El-Boghdadly K et al. Risks to healthcare workers following tracheal intubation of patients with COVID-19: a prospective international multicentre cohort study**.

Reason for exclusion: wrong study design: descriptive study

**Xinghuan W et al. Association between 2019-nCoV transmission and N95 respirator use**.

Reason for exclusion: wrong study design: quasi-experimental study

**Clariot S et al. Minimising COVID-19 exposure during tracheal intubation by using a transparent plastic box: a randomised prospective simulation study**.

Reason for exclusion: wrong study design: a simulation study

**Schumacher J et al. The impact of respiratory protective equipment on difficult airway management: a randomised, crossover, simulation study**.

Reason for exclusion: wrong study design: a simulation study

**Gong Y et al. Anesthesia Considerations and Infection Precautions for Trauma and Acute Care Cases During the COVID-19 Pandemic: Recommendations From a Task Force of the Chinese Society of Anesthesiology**.

Reason for exclusion: wrong study design: clinical recommendation

**Appendix Table I:** Systematic Reviews.

| Study | Review objectives | Descriptions of interventions/phenomena of interest | Descriptions of outcomes included in the review | Descriptions of contexts included in the review | Search details | Number of studies and participants included | Appraisal instruments used |
| --- | --- | --- | --- | --- | --- | --- | --- |
| Abdelrahman T, et al. 2020. | To identify and collate global experience, practice and recommendations relating to OR practice in the context of the COVID-19 pandemic | Specific practices, or providing recommendations or guidance relating to OR practice, in the context of the COVID-19 pandemic | Five domains for data capture were identified a priori: 1) the physical OR factors, 2) personnel factors, 3) patient factors, 4) procedure factors, 5) other considerations. | Healthcare settings amidst COVID-19 pandemic | Articles reporting specific practices, experience, recommendations, or guidance in relation to emergency OR practice, in the context of the COVID-19 pandemic, were included. Articles were excluded if they did not meet the inclusion criteria, if the full-text article was unavailable, or if no English version was extractable. | 11 studies. two of which had cohorts of four and 17 | Oxford Centre for Evidence-Based Medicine (OCEBM) |
| Ana L, Andrew JS, Rhonda S. 2020. | To evaluate the effect of powered air-purifying respirators (PAPRs) as part of respiratory protection versus another device (N95/FFP2) on HCW infection rates and contamination | Powered Air-Purifying Respirator (PAPR) studied separately or within Personal Protective equipment vs. Any other respiratory protective equipment, FFP3/FFP2/N95 or surgical masks | 1) healthcare worker infection rates utilising PAPR technology within a PPE program as infection with SARS-Cov-2, SARS-Cov-1, EBOLA or MERS; 2) contamination of skin or clothing measured with any type of test material to visualise contamination; 3) compliance with guidance on use of PAPR measured with e.g. observation checklist; 4) level of wearer comfort whilst using the PAPR; - objective and/or subjective measures of work of breathing during the use of PAPR; 5) costs of resource use of PAPR equipment; 6) impact of structured training programs on PAPR use | Healthcare settings amidst COVID-19 pandemic | P: Healthcare workers Volunteers I: Powered Air-Purifying Respirator (PAPR) studied separately or within Personal Protective equipment C: Any other respiratory protective equipment, FFP3/FFP2/N95 or surgical masks O: - healthcare worker infection rates utilising PAPR technology within a PPE program as infection with SARS-Cov-2, SARS-Cov-1, EBOLA or MERS; - contamination of skin or clothing measured with any type of test material to visualise contamination; - compliance with guidance on use of PAPR measured with e.g. observation checklist; - level of wearer comfort whilst using the PAPR; - objective and/or subjective measures of work of breathing during the use of PAPR; - costs of resource use of PAPR equipment; - impact of structured training programs on PAPR use S: Randomised controlled trials, Non-randomised studies, Observational studies (cohort studies, case-control studies, cross-sectional studies, case series) S: Inpatient care/Critical Care/Intensive Care, Perioperative process-preadmission, preoperative, intraoperative and postoperative setting Timing: Perioperative process-preadmission, preoperative, intraoperative and postoperative setting | Ten eligible studies. Five of these studies were simulation studies. Two of the studies were randomised controlled trials. A single study was a randomised controlled trial in a simulation setting. A single study was a randomised controlled trial in a simulation setting. A single study was an observational case series of healthcare workers (airway proceduralist only) managing patients infected with SARS-CoV-2 in China at the start of 2020. Two were observational studies with control cohorts. One observational simulation study was a case series without a control group. | Cochrane RoB and ROBINS-I |
| Bartoszko JJ, Farooqi MAM, Alhazzani W, Loeb M. 2020. | To compare medical masks to N95 respirators in preventing laboratory-confirmed viral infection and respiratory illness including coronavirus specifically in healthcare workers. | N95 vs medical masks | Viral respiratory infection laboratory confirmed by PCR, serology, or viral culture (our primary outcome), laboratory-confirmed coronavirus infection, laboratory-confirmed influenza, infection, influenza-like illness, clinical respiratory illness, or workplace absenteeism. | Healthcare settings amidst COVID-19 pandemic | P: healthcare workers (defined as workers in a healthcare setting that could be exposed to a patient with acute respiratory illness) I: medical masks (defined surgical, procedural, isolation, laser, fluid resistant or face masks certified for use as a medical device) C: N95 respirators (defined as respirators were N95 filtering face piece respirators certified by the National Institute for Occupational Safety and Health (NIOSH) and European standard filtering face-piece (FFP2) respirators) O: viral respiratory infection laboratory confirmed by PCR, serology, or viral culture (our primary outcome), laboratory-confirmed coronavirus infection, laboratory-confirmed influenza infection, influenza-like illness, clinical respiratory illness, or workplace absenteeism | Four RCTs | Modified Cochrane RoB |
| Carl-Etienne J, Toma P, Matt B, Genevieve G, Louise P. 2020. | To provide a comprehensive summary of the evidence on pandemic control, with a focus on cost-effectiveness. | Effectiveness of hand washing and face masks. | cost-effectiveness | Healthcare settings amidst COVID-19 pandemic | Articles reporting on the effectiveness or cost-effectiveness of at least one intervention were included and grouped into higher-quality evidence (randomised trials) and lower-quality evidence (other study designs). | 62 studies | Did not examine risk of bias. |
| Chou R, Dana T, Jungbauer R, Weeks C, McDonagh MS. 2020. | To evaluate the effectiveness of respirators (e.g., N95) versus facemasks (surgical) versus cloth masks for prevention of COVID-19 in addition to standard precautions (gowns + gloves + handwashing) in community settings, high-risk healthcare settings (e.g., intensive care unit, emergency department), and in healthcare settings with close contact but unknown risk (e.g., primary care, other settings) To also evaluate the evidence for extended use or reuse of N95 respirators for prevention of COVID-19. | N95 respirators or equivalent, surgical/medical masks, and cloth masks. | 1) Infection with SARS-CoV-2, SARS-CoV-1, or MERS-CoV 2) Influenza-like illness, lab-confirmed viral infection, lab-confirmed influenza, and clinical respiratory illness 3) Harms of mask usage | Healthcare settings amidst COVID-19 pandemic | Healthcare workers or community members at risk of contracting COVID-19 or other viral respiratory illnesses due to workplace or community-based exposure. Randomised controlled trials of one mask type versus another for prevention of COVID-19 (the disease caused by SARS-CoV-2 infection), SARS-1 (the disease caused by SARS-CoV-1 infection), and MERS (the disease caused by MERS-CoV infection). influenza-like illness, and laboratory-confirmed viral respiratory illness. Randomised controlled trials of masks versus no masks (to inform indirect comparisons) for prevention of COVID-19, SARS-1, MERS, influenza-like illness, and laboratory-confirmed viral respiratory illness. Cohort and case-control studies on effects of mask use and risk for prevention of COVID-19 (the disease caused by SARS-CoV-2 infection), SARS-1 (the disease caused by SARS-CoV-1 infection), and MERS (the disease caused by MERS-CoV infection). Randomised controlled trials, cohort studies, and case-control studies on reuse or extended use of masks versus standard use for prevention of COVID-19, SARS-1, or MERS. | 39 studies (18 randomised controlled trials and 21 observational studies; 33 867 participants) | Criteria adapted from the US Preventive Services Task Force (USPSTF). |
| Chu DK, Akl EA, Duda S, Solo K, Yaacoub S, Schünemann HJ, et al. 2020. | To provide a quantitative assessment on the physical distance associated with reduced risk of acquiring infection when caring for an individual infected with SARS-CoV-2, SARS-CoV, or MERS-CoV. | optimum distance for avoiding person-to-person virus transmission and to assess the use of face masks and eye protection to prevent transmission of viruses. | 1) risk of transmission to people in healthcare or non-healthcare settings by those infected; 2) contextual factors such as acceptability, feasibility, effect on equity, and resource considerations related to the interventions of interest. | Healthcare settings amidst COVID-19 pandemic | Studies of any design and in any setting that included patients with WHO-defined confirmed or probable COVID-19, SARS, or MERS, and people in close contact with them, comparing distances between people and COVID-19 infected patients of 1 m or larger with smaller distances, with or without a face mask on the patient, or with or without a face mask, eye protection, or both on the exposed individual. | 172 observational studies and 44 relevant comparative studies (n=25,697 patients) | Standardised prepiloted forms (covidence); cross-checked screening results using artificial intelligence (Evidence Prime) |
| Elaine T, Yvonne C, Christopher B, Simon S, Michael S, Xin-Hui C, et al. 2020. | To summarise recommendations regarding the extended use or reuse of N95 respirators, and to provide an overview of published information by regulatory authorities, surveillance organisations, and ministries of health of several countries. | N95 Respirators | N/A | Healthcare settings amidst COVID-19 pandemic | Two researchers independently scrutinised the websites of the regulatory authorities of countries or regions and of ministries of health that a members or associates of the International Coalition of Medicines Regulatory Authorities (ICMRA). | N/A | N/A |
| Fouladi Dehaghi B, Ghodrati Torbati A, Teimori G, Ghavamabad i LI, Jamshidnezhad A. 2020. | To investigate the effectiveness of face masks against respiratory infections, including coronavirus. | Respirator and non-respirator masks | settings, mask type, findings | Healthcare settings amidst COVID-19 pandemic | A survey of articles published up to Apr 30 2020 about the effectiveness of face masks against coronavirus infections was performed using four electronic databases: PubMed, Scopus, Science Citation Index (Web of Science), and Google scholar. The following terms were used in the search strategy: ['Facemask' OR 'Facemasks' OR 'Mask' OR 'Masks' OR 'Respirator' OR 'Respirators'] AND ['COVID-19' OR 'Coronavirus' OR 'Novel Coronavirus'] AND ['Medical staff' OR 'Healthcare staff' OR 'health care providers' OR 'Healthcare providers' OR 'Healthcare workers' OR 'Health workers' OR 'Healthcare Professionals']. | 5 studies | N/A |
| Katie O, Gertsman S, Sampson M, Webster R, Tsampalieros A, Ng R, et al. 2020. | To synthesise existing data on the effectiveness of ultraviolet germicidal irradiation (UVGI) for N95 FFR decontamination. | decontamination procedures for N95 (including SN95) filtering facemask respirators or their components | 1) effectiveness of the UVGI method at removing viral or bacterial load; 2) impact of the UVGI method on filtering face mask performance, with a specific focus on aerosol penetration and airflow resistance (pressure drop); or 3) measures or observations related to change in mask fit or physical degradation of the mask following UVGI exposure. | Healthcare settings amidst COVID-19 pandemic | Original research reporting on function, decontamination, or mask fit following UVGI were included. | Thirteen studies were identified, comprising 54 UVGI intervention arms and 58 N95 models. | Independent instrument evaluating population, design, methodology , outcome |
| Liang M, Gao L, Cheng C, Zhou Q, Uy JP, Heiner K, et al. 2020. | To evaluate the effectiveness of the use of masks to prevent laboratory-confirmed respiratory virus transmission. | Protective effect of masks against spread of respiratory infections | protective effect of mask vs respiratory viral infections | Healthcare settings amidst COVID-19 pandemic | The studies meeting the following criteria were included: (1) concerning the relationship between the face mask and preventing RVIs; (2) diagnosis of respiratory virus must have laboratory evidence, or the local clinical diagnostic criteria are applied during an acute large-scale infectious disease when laboratory evidence might be not available; (3) complete data available of both cases and controls to calculate an odds ratio (OR) with 95% confidence interval (CI); (4) appropriate study design; (5) no language restrictions applied. The exclusive criteria were as follows: (1) conferences/meetings abstracts, case reports, editorials, and review articles; (2) duplicate publication or overlapping studies; (3) studies with unavailable full texts. | 21 studies which included 13 case-control studies, 6 cluster randomised trials, and 2 cohort studies. 12 studies investigated HCWs. 7 studies investigated Severe acute respiratory syndrome coronavirus (SARS-CoV), 12 studies investigated influenza virus including 5 investigating the H1N1 virus, and 1 study investigated SARS-CoV-2. | The Newcastle-Ottawa Scale (NOS) and the Jadad scale. |
| MacIntyre CR, Chughtai AA. 2020. | To review the evidence around the efficacy of masks and respirators for healthcare workers, sick patients and the general public. | Use of respiratory protection by healthcare workers, sick patients and community members | respiratory viral infection | Healthcare settings amidst COVID-19 pandemic | Randomised controlled trials on masks and respirators. Studies that were not randomised controlled trials, were about anesthesia, or not about prevention of infection were excluded. Animal studies, experimental and observational epidemiologic studies were also excluded. Studies published in English language were included | 19 studies | N/A |
| Offeddu V, Yung CF, Low MSF, Tam CC. 2017. | To develop evidence-based recommendations to reduce the occupational risk of respiratory infection among medical personnel. | Protective effect of facemasks and respirators against respiratory infections among healthcare workers | Effectiveness of PPE in reducing the risk of clinical or laboratory-confirmed respiratory outcomes; | Healthcare settings amidst COVID-19 pandemic | P: HCWs; I: Any type of PPE; C: no PPE O: Effectiveness of PPE in reducing the risk of clinical or laboratory-confirmed respiratory outcomes; S Healthcare settings worldwide. Study design: Published, peer-reviewed randomised control trials (RCTs) and observational studies; | 6 RCTs and 23 observational studies | Cochrane RoB and RevMan for RCTs. Adapted Newcastle-Ottawa scales for case-control, cohort, and cross-sectional studies. |
| Prashanth R, Jonathan Thomas S, Ruben D, Christopher A, Shehan H. 2020. | To evaluate the evidence behind UK Government PPE Guidance on surgical masks versus respirators for SARS-CoV-2 protection in patient-facing healthcare workers delivering secondary care. | UK guidelines comparing surgical facemasks and respirators specific to SARS-CoV-2 | For laboratory studies, these data included study design, research question, masks/respirators tests, testing particle/pathogen, findings, and appraisal comments. For clinical studies, these data included setting, participants, interventions, outcomes, and limitations raised in appraisal. | Healthcare settings amidst COVID-19 pandemic | Any comparison between FRSM and respirator. Any study design containing primary data | 9 studies | CASP Checklists |
| Tom J, Mark J, Lubna AAA, Ghada B, Elaine B, Justin C, et al. 2020. | To examine the effectiveness of eye protection, face masks, or person distancing on interrupting or reducing the spread of respiratory viruses. | Social distancing, mask and hand hygiene either as stand-alone interventions or combined | any acute respiratory illness and its related consequences | Healthcare settings amidst COVID-19 pandemic | People of any age using face masks (i.e. surgical or medical masks and N95 respirators), eye protection, or person distancing against standard practice, or a similar physical barrier, or compared any of these interventions. Studies that reported a measure of acute respiratory illness – such as influenza-like illness, influenza, or respiratory infections – and/or its consequences (e.g. days off work, complications, hospitalisation and death, if clearly reported as consequences of the respiratory illness). | 15 randomised trials investigating the effect of masks (14 trials) in healthcare workers and the general population and of quarantine (1 trial). | Cochrane RoB |
| Verbeek JH, Rajamaki B, Ijaz S, Sauni R, Toomey E, Blackwood B, et al. 2020. | To evaluate which type of full-body PPE and which method of donning or doffing PPE have the least risk of contamination or infection for HCW, and which training methods increase compliance with PPE protocols. | PPE with PAPR, more protective attire, not permeable not breathable (A), gowns versus aprons, one type of full-body PPE, sealed gown-glove interface, gown: easy to doff, gown with gown-glove improvement, gloves with tab, mask tabs, CDC method of doffing, single-step doffing, doffing with double gloves, donning and doffing with instructions, doffing with extra sanitation of gloves, doffing with hypochlorite, video-based learning | 1) contamination of skin or clothing, measured with any type of test material to visualise contamination (e.g. stains made visible with UV light) or harmless viruses or bacteria; 2) infection with EVD, another viral haemorrhagic fever, or comparable highly infectious disease with serious consequences such as SARS, or COVID-19; 3) compliance with guidance on selection of type and use of PPE measured, for example, with an observation checklist 4) User-reported assessment of comfort and convenience 5) Costs or resource use 6) Time to don and doff the PPE. | Healthcare settings amidst COVID-19 pandemic | Controlled studies that evaluated the effect of full-body PPE used by HCW exposed to highly infectious diseases, on the risk of infection, contamination, or non-compliance with protocols. Studies that compared the effect of various ways of donning or doffing PPE, and the effects of training on the same outcomes. | 24 studies with 2278 participants, of which 14 were randomised controlled trials (RCT), one was a quasi-RCT and nine had a non-randomised design. Eight studies compared types of PPE. Six studies evaluated adapted PPE. Eight studies compared donning and doffing processes and three studies evaluated types of training. Eighteen studies used simulated exposure with fluorescent markers or harmless microbes. | Cochrane RoB and ROBINS-I |
| Zorko DJ, Gertsman S, O'Hearn K, Timmerman N, Ambu-Ali N, Dinh T, et al. 2020. | To identify and synthesise data from original research evaluating interventions to decontaminate surgical masks for the purpose of reuse | dry heat, autoclave, ethanol, isopropanol, bleach | mask performance (filtration efficiency [FE] and airflow resistance); reduction in pathogen load; in vivo infection rates following use of treated masks; mask appearance or physical degradation; or adverse effects experienced by the wearer. | Healthcare settings amidst COVID-19 pandemic | P: HCWs donning surgical facemask PPE or their components<br>I: intervention(s) to decontaminate, sterilise or treat surgical masks (applied either before or after their use) for the purposes of reuse as PPE<br>C: no decontamination<br>O: a) mask performance (i.e. filtration efficiency and airflow resistance); b) reduction in pathogen load; c) in vivo infection rates following use of decontaminated masks; d) changes in physical appearance (i.e. mask appearance or physical degradation); e) adverse effects experienced by the wearer (e.g. skin irritation); or f) feasibility of the intervention (e.g. time, cost, resource utilisation). Prospective original research on decontamination interventions for surgical masks. Original research, including systematic reviews | 7 studies.<br>Decontamination (n=1)<br>Pre-contamination (n=6) | Independent instrument evaluating RoB in study design, methodological consistency, population heterogeneity, sampling bias, outcome evaluation, and selective reporting |
| Iannone P, Castellini G, Coclite D, Napoletano A, Fauci AJ, Iacorossi L, et al. 2020. | To assess the role of N95 respirators in protecting HCWs during the COVID-19 pandemic | N95 respirators versus surgical masks. | Primary outcomes: 1) SARS-CoV-2 infection; 2) Clinical respiratory illness (CRI).<br>Secondary outcomes: (3) Influenza like illness (ILI); (4) Laboratory-confirmed respiratory viral infection; (5) Laboratory-confirmed bacterial colonisation; (6) Laboratory-confirmed respiratory infection; (7) Laboratory-confirmed influenza; (8) Discomfort of wearing respiratory protections. | Healthcare settings amidst COVID-19 pandemic | Randomised controlled trials (RCTs) run in healthcare settings. Randomisation was allowed both at individual and cluster level. | Four RCTs involving 8736 HCWs. No trial specifically on prevention of COVID-19 | Cochrane RoB |

**Appendix Table II:** Case Series.

| Study | Country | Setting/context | Participant characteristics | Groups | Outcomes measured |
| --- | --- | --- | --- | --- | --- |
| Wilson JM, Schwartz AM, Farley KX, Devito DP, Fletcher ND. 2020. | USA | infectious complications, perioperative bodily fluid occupational exposure | 1,270 patients who underwent CRPP of a supracondylar humeral fracture and met inclusion criteria were identified. Patients had a mean age of 6 years, and sex approached an even divide between male patients and female patients. Over 90% of patients stayed in the hospital <24 hours. The proportion of fracture occurrence and treatment for the entire cohort was greatest in April to June (30.2%), followed by July to September (27.0%), October to December (25.3%), and January to March (17.5%). | Sterile (n=874)Semi-Sterile (n=396) | 1 of 2 large, metropolitan, pediatric hospitals within the same hospital system. |

**Appendix Table III:**
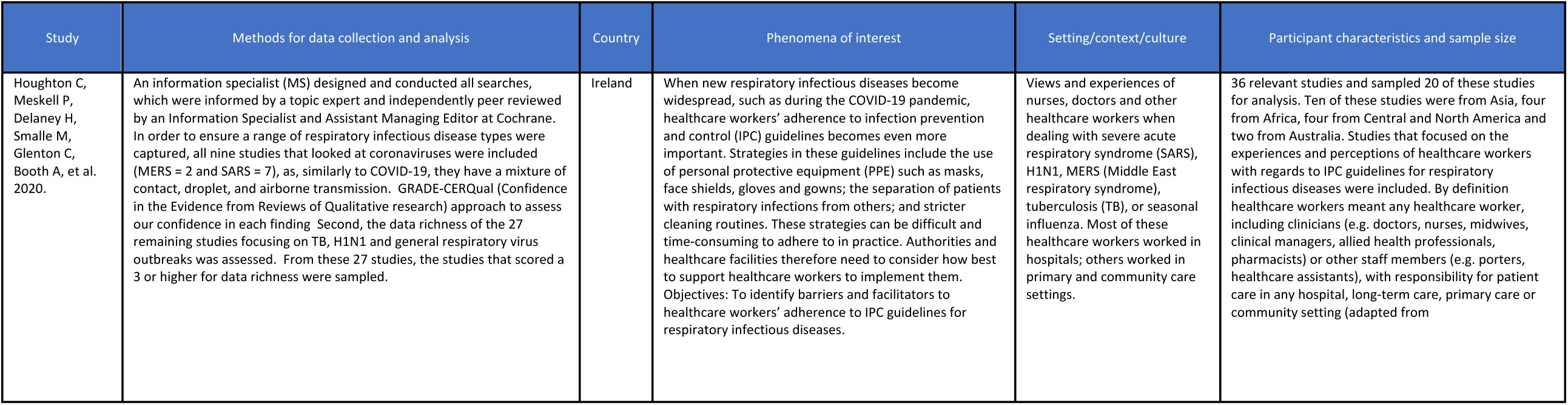
Qualitative Research.

